# Targeting anaemia without measuring it: surrogate prediction, district decision uncertainty and the value of repeat measurement in India

**DOI:** 10.64898/2026.08.04.26359555

**Authors:** H S Siddalingaiah

## Abstract

**Background & objectives:** India’s fifth National Family Health Survey measured anaemia in all 707 districts; the sixth did not. Anaemia now sits in a venous-blood survey covering 183 districts that reports nationally, not by district. Using the last round that measured it, we asked whether the surviving indicators reproduce it, whether losing it changes which districts a programme prioritises, and what repeat measurement is worth.

**Methods:** We estimated district anaemia and its uncertainty for children aged 6–59 months and non-pregnant women aged 15–49 years with a small-area model using design-based variances; tested prediction from the retained indicators on districts and whole States never seen; compared prioritisation under three information sets with matched preference draws; and valued a repeat measurement of the same latent prevalence.

**Results:** District estimates carried a median standard error of 3.57 percentage points (pp) for children and 2.22 pp for women. The best surrogate had a root mean squared error of 10.14 pp for children, of which 9.44 pp was structural — a ratio of 2.5 to the root-mean-square measurement error — and in unseen States did worse than the training mean. Of 71 districts selected on current estimates, 19.1 fall outside the latent top 71. Measuring 183 districts recovered 46.3% of that gap when chosen for decision value, against 9.6% under equal allocation across States.

**Interpretation & conclusions:** Input coverage cannot stand in for measured anaemia. Where measurement is bounded, which districts are measured matters more than how many, provided the platform difference is bridged first.

## Introduction

For three decades every round of the National Family Health Survey measured anaemia. The fifth round (NFHS-5, 2019–21) found it in 67.1% of children aged 6–59 months and 57.0% of women aged 15–49 years, from finger-prick samples in 707 districts,^1^ and those numbers have driven programme design ever since. The sixth round (NFHS-6, 2023–24) reports no anaemia at all.^2^ This was not an omission at publication: the blood was never drawn. Officials have been reported as saying the finger-prick method was judged unreliable and prone to overstating anaemia, and that the task would pass to a venous-blood survey; no primary departmental statement appears to have been published.^3^ The reasoning is sound, since current guidance prefers venous blood on an automated analyser and holds that finger-prick values cannot simply be converted into venous ones.^4^

That successor is the Survey for Assessment of Markers of Population Health, Activity, Diet and Anthropometry, first known as the Diet and Biomarkers Survey in India: about two lakh participants in 183 districts across 35 States and Union Territories.^5^ It draws venous samples,^6^ and exists for national and State estimation, the level it reports at. India has swapped reach for accuracy, and no current district figure for anaemia now exists anywhere.

The timing matters. On 29 June 2026 the Ministry of Health and Family Welfare released the Anaemia Mukt Bharat Abhiyaan guidelines, organising the programme around four verbs — Test, Treat, Talk and Track.^7^ A programme built around tracking must now work, in most districts, with no current measure of the thing it exists to change. What survives is the record of the interventions: iron and folic acid in pregnancy, up from 44.1% to 54.9%, alongside antenatal care, immunisation and infant feeding.^2^

Comment so far has been about transparency. The prior question is practical, because a measurement earns its cost only when it changes a decision.^8,9^ We put three questions to the last round that measured district anaemia. Can the surviving indicators reproduce it? Does losing it change which districts would be prioritised? And what would repeat measurement be worth, and where?

## Methods

The four analytic steps are detailed in full in the supplementary material.

### Study design, setting and data sources

This is a cross-sectional secondary analysis of Indian districts; the unit of analysis is the district. No participant was contacted, and de-identified records were used only to compute sampling variances. Reporting follows STROBE.^10^

Three sources were used. The NFHS-5 district fact sheet (2019–21) supplied anaemia prevalence and the ten prioritisation criteria; these are the published numbers the programme works from, and so are the point estimates. The NFHS-5 person recode supplied what fact sheets never print — how precisely each figure is known — from 1 685 994 people with a valid haemoglobin reading, with standard errors following the survey’s two-stage design of weights, strata and clusters.^11^ The NFHS-6 fact sheets supplied the surviving indicators, defining the reduced information set of Step 3. Taking estimate and precision from different routes is justified by their agreement, reported in the supplement.

Two populations were treated equally: children aged 6–59 months (below 11.0 g/dl) and non-pregnant women aged 15–49 years (below 12.0 g/dl), the survey’s own operational definitions, retained because the estimand is the published district figure rather than a reestimate of prevalence; the age-specific child cut-offs of current guidance^4^ are tested separately. Pregnant women were excluded because only 21.4% of districts hold a reliably reported value. Anaemia is never compared across rounds, the measurement not being established as comparable across platforms.^4^

### Step 1. District anaemia and its uncertainty

A district figure resting on a median of 243 measured children (Table Ia) is too shaky to treat as fact. Each was modelled on the logit scale as a noisy reading of an underlying value, districts nested inside States, in a Fay–Herriot model^12,13^ that pulls a thinly measured district towards its State and leaves a well-measured one alone. The observation variance was fixed at the design-based value; variance components carried half-Cauchy priors.^14^ Four Gibbs chains of 4000 retained draws followed 1000 burn-in iterations. Convergence was judged by the potential scale reduction factor for every district and variance component, by effective sample sizes, by a posterior predictive check, and by refitting under different priors (S1.3).

**Table I.**
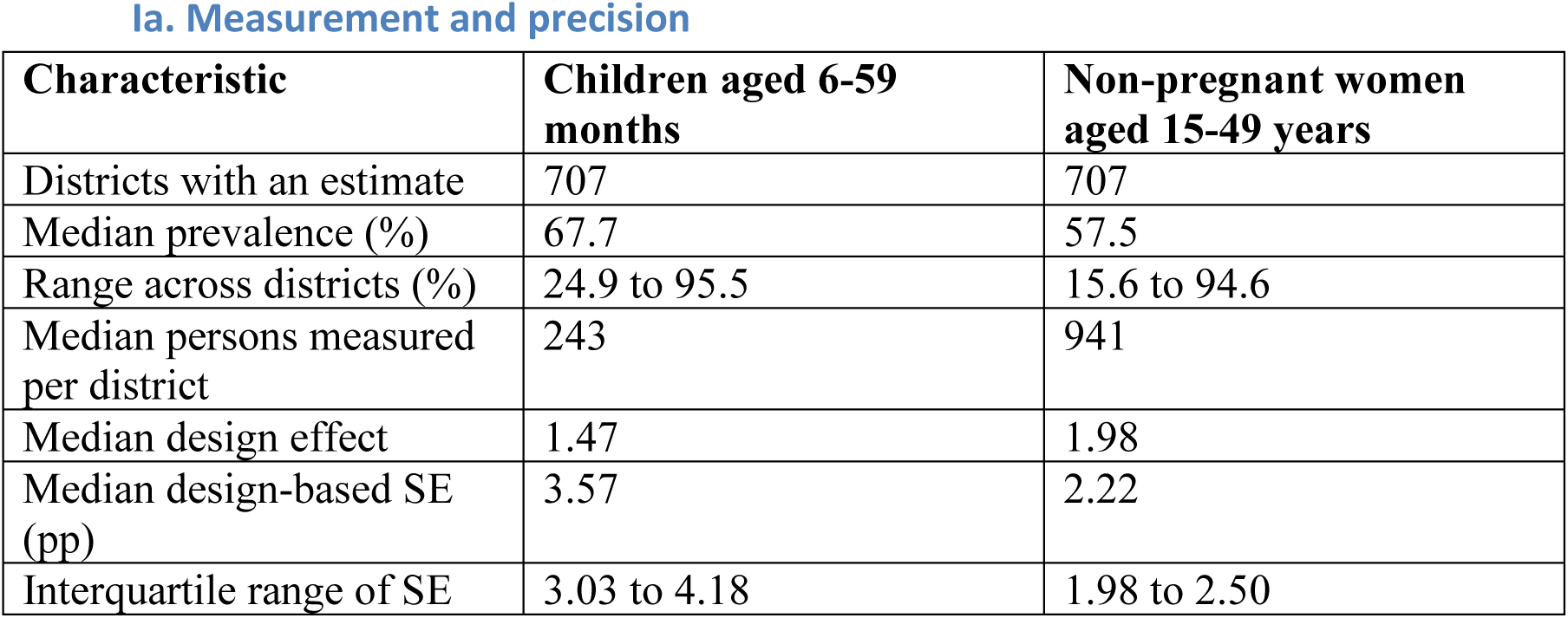
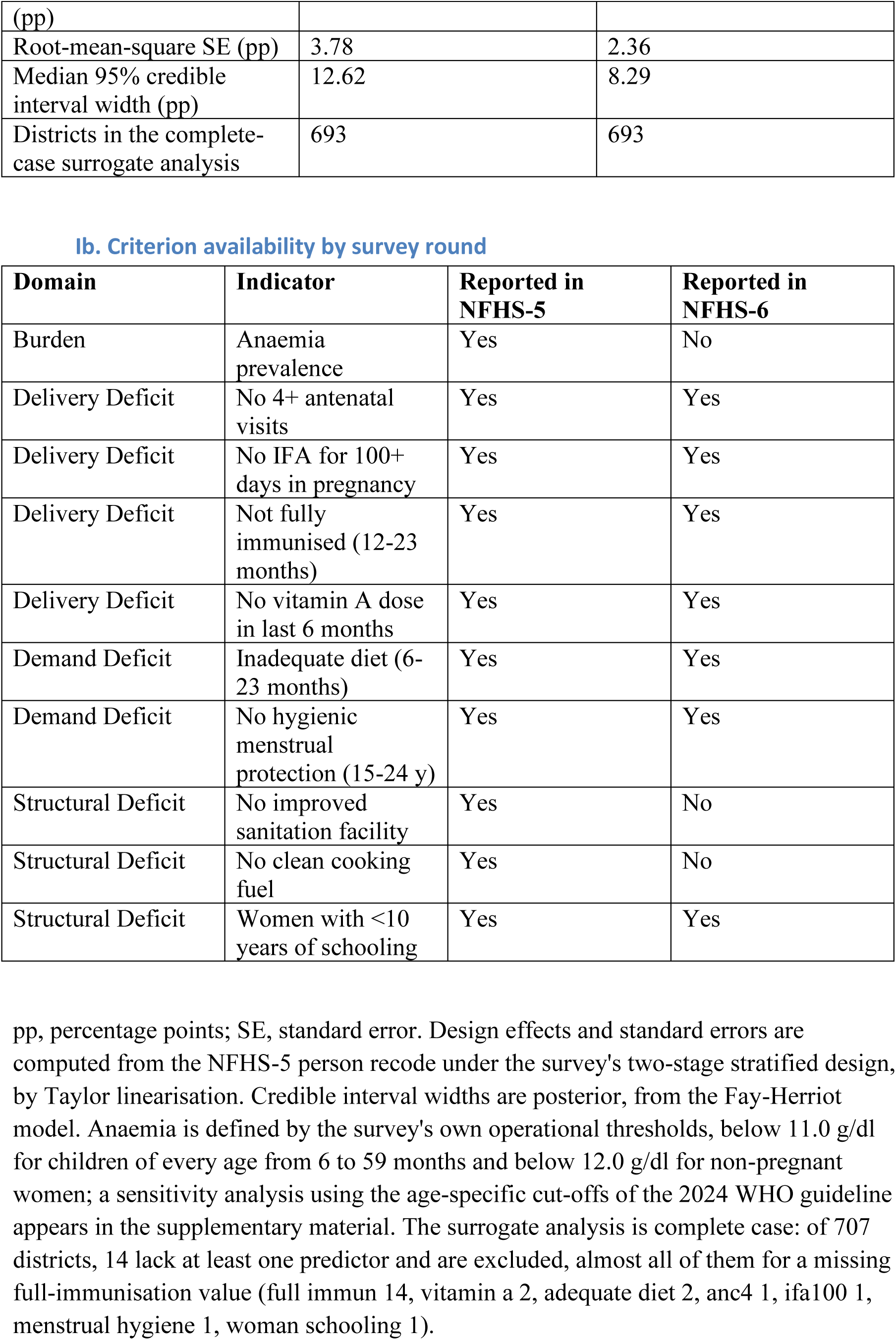
District anaemia measurement in NFHS-5, and the availability of each prioritisation criterion in NFHS-6.

### Step 2. Can the surviving indicators substitute?

If district anaemia could be predicted from the indicators NFHS-6 still reports, its removal would cost little. We tested that with a mean-only baseline refitted within each training fold, a penalised linear model, and gradient-boosted trees.^15^ Random ten-fold cross-validation asks how well a model fills gaps among districts whose neighbours it has seen; grouped cross-validation, hiding every district of a State at once, asks the harder question a real surrogate would face. A second predictor set added every other NFHS-5 indicator, so a poor result could not be blamed on withheld information. The analysis is complete case on 693 of 707 districts, the exclusions being almost all districts lacking a published full-immunisation value.

Because the outcome is itself measured with error, we subtracted its sampling variance from the observed mean squared error, leaving the structural part attributable to the model, and compared that with the root-mean-square district standard error rather than a median (S2.3). Calibration slopes are reported, since a model can post a respectable error while compressing predictions towards the mean. This step was not planned in advance: it replaced an efficiency model that failed to identify (S7), which carries no weight here. The replacement is exploratory, and the whole-State test was adopted as the tougher standard for that reason.

Associations between each surviving input and anaemia were also estimated with State fixed effects and robust standard errors, with a Holm adjustment across the seven indicators, then applied to the national gains between rounds through the full covariance matrix of the coefficients. These are ecological; the projection is descriptive, not a forecast.

### Step 3. Does the loss change which districts are chosen?

Districts were ranked by TOPSIS^16^ on ten criteria in four domains — burden, delivery gaps, demand gaps and structural disadvantage — each oriented so a higher value means a stronger claim on attention, and listed in Table Ib. Stochastic multicriteria acceptability analysis^17,18^ ran 10 000 rounds, each drawing criterion values from the Step 1 posteriors and a weight vector at random, ranking all districts, and recording which reached the top tenth. Weights were drawn hierarchically, across domains and then within each, so a domain does not gain weight merely by containing more indicators. The three information sets — all NFHS-5 criteria, only those NFHS-6 retains, and the NFHS-5 criteria minus anaemia — were compared under matched preference draws, removing the weight of an unavailable criterion and renormalising the rest rather than redrawing. Districts are reported as classes, never ranks: robust at 80% of rounds or more, conditional at 20–80%, otherwise not prioritised. Class boundaries and the weight model were varied.

### Step 4. What the uncertainty costs

This decision is simpler than Step 3 and separate from it: pick the 71 districts with the highest burden, benefit being proportional to true prevalence. The quantity reported is expected non-membership — of the 71 districts chosen on posterior means, how many fall outside the latent top 71 in a posterior draw. It is a property of the posterior, not a count of observed mistakes, and says nothing about the multicriteria rule.

The measurement valued is a repeat observation of that same 2019–21 latent prevalence, on the same platform, each district carrying its own design-based sampling error, with belief updated on the logit scale by precision weighting. It is not an estimate of what any survey decision has cost, and does not evaluate the successor survey’s district list, which is not public. Eight rules were scored, four using the decision and four ignoring it: random selection, equal allocation across States and Union Territories, and allocation proportional to district count and to measured population. Districts were chosen on one half of the draws and scored on the other, the ordering statistic included, and every rule was scored 40 times with a fresh frame where it needs one and fresh survey outcomes throughout. Networks of 25 to 300 districts were tested; sensitivity analyses relaxed stationarity, allowed a platform difference, and varied achieved sample and decision size.

Analyses used Python 3.14 under a fixed seed. An AI coding assistant (Claude, Anthropic) helped write and debug the analysis code and draft and edit manuscript text; it was not used to generate, select or interpret any result. Every number reported here comes from the analysis code and is re-checked against its output by a verification script supplied with the submission.

## Results

### Step 1. How precisely anaemia was known

The median district contributed 243 measured children and 941 measured women. Median design effects were 1.47 and 1.98, and median design-based standard errors 3.57 pp and 2.22 pp, with the middle half of child estimates between 3.03 and 4.18 pp. After the small-area model, the median district’s 95% credible interval was 12.62 pp wide for children and 8.29 pp for women (Table Ia): at best, known to within about six percentage points either way. All district-level potential scale reduction factors were at most 1.0005, the variance components mixed freely, and refitting under wider and narrower priors moved no district posterior mean by more than 0.35 pp.

### Step 2. The surviving indicators cannot reproduce the missing one

Gradient-boosted trees, given every indicator NFHS-6 keeps, predicted district child anaemia with a root mean squared error of 10.14 pp, a mean absolute error of 7.85 pp, and out-of-sample R^2^ of 0.270. Removing the outcome’s own sampling variance leaves a structural error of 9.44 pp against a root-mean-square measurement error of 3.72 pp, a ratio of 2.5; for women, 9.80 pp against 2.34 pp, a ratio of 4.2 (Table II). Predictions were also compressed towards the mean: calibration slopes were 0.76 for children and 0.80 for women.

**Table II.** Out-of-sample accuracy of surrogate models for district anaemia prevalence.

| Population | Predictor set | Validation | Penalised linear | Boosted trees |
| --- | --- | --- | --- | --- |
| Children | NFHS-6 retained | random folds | 0.239 (10.35) | 0.270 (10.14) |
| Children | NFHS-6 retained | held-out states | 0.173 (10.79) | −0.069 (12.27) |
| Children | All NFHS-5 | random folds | 0.353 (9.55) | 0.450 (8.81) |
| Children | All NFHS-5 | held-out states | 0.198 (10.63) | 0.136 (11.04) |
| Women | NFHS-6 retained | random folds | 0.185 (10.84) | 0.295 (10.08) |
| Women | NFHS-6 retained | held-out states | 0.009 (11.95) | −0.355 (13.98) |
| Women | All NFHS-5 | random folds | 0.290 (10.11) | 0.528 (8.25) |
| Women | All NFHS-5 | held-out states | −0.089 (12.53) | −0.055 (12.33) |

Cells give the out-of-sample $R^2$ with the root mean squared error (RMSE) in percentage points in parentheses. A negative $R^2$ means the model predicts worse than the mean of its own training folds. Random folds are ten-fold cross-validation over districts; held-out states withhold every district of a state, so no district of the target state is seen in training. The published district estimate used as the outcome is itself measured with error, so the RMSE contains both the model's error and the survey's: for children, the mean-only baseline reached $R^2$ $-0.001$ and $-0.023$ , the best model's mean absolute error was 7.85 pp, and after removing the sampling variance of the outcome its structural error was 9.44 pp against a root-mean-square district standard error of 3.72 pp, a ratio of 2.5; for women, the mean-only baseline reached $R^2$ $-0.001$ and $-0.050$ , the best model's mean absolute error was 7.96 pp, and after removing the sampling variance of the outcome its structural error was 9.80 pp against a root-mean-square district standard error of 2.34 pp, a ratio of 4.2. The structural error and the standard error are both root-mean-square quantities and are therefore directly comparable.

Sent to a State it had never seen, the flexible model broke: R^2^ fell to −0.069 for children and −0.355 for women, against −0.023 and −0.050 for a mean-only baseline, and calibration slopes to 0.42 and 0.11 (Fig. 1). Every model is penalised by this test, the flexible one most; the best traveller was the plain linear model, and even it reached only 0.173 for children and 0.009 for women (Table II). Every other NFHS-5 indicator lifted random-fold R^2^ to 0.450 and 0.528, yet for women nothing then reached an unseen State better than the training mean.

**Figure 1.**
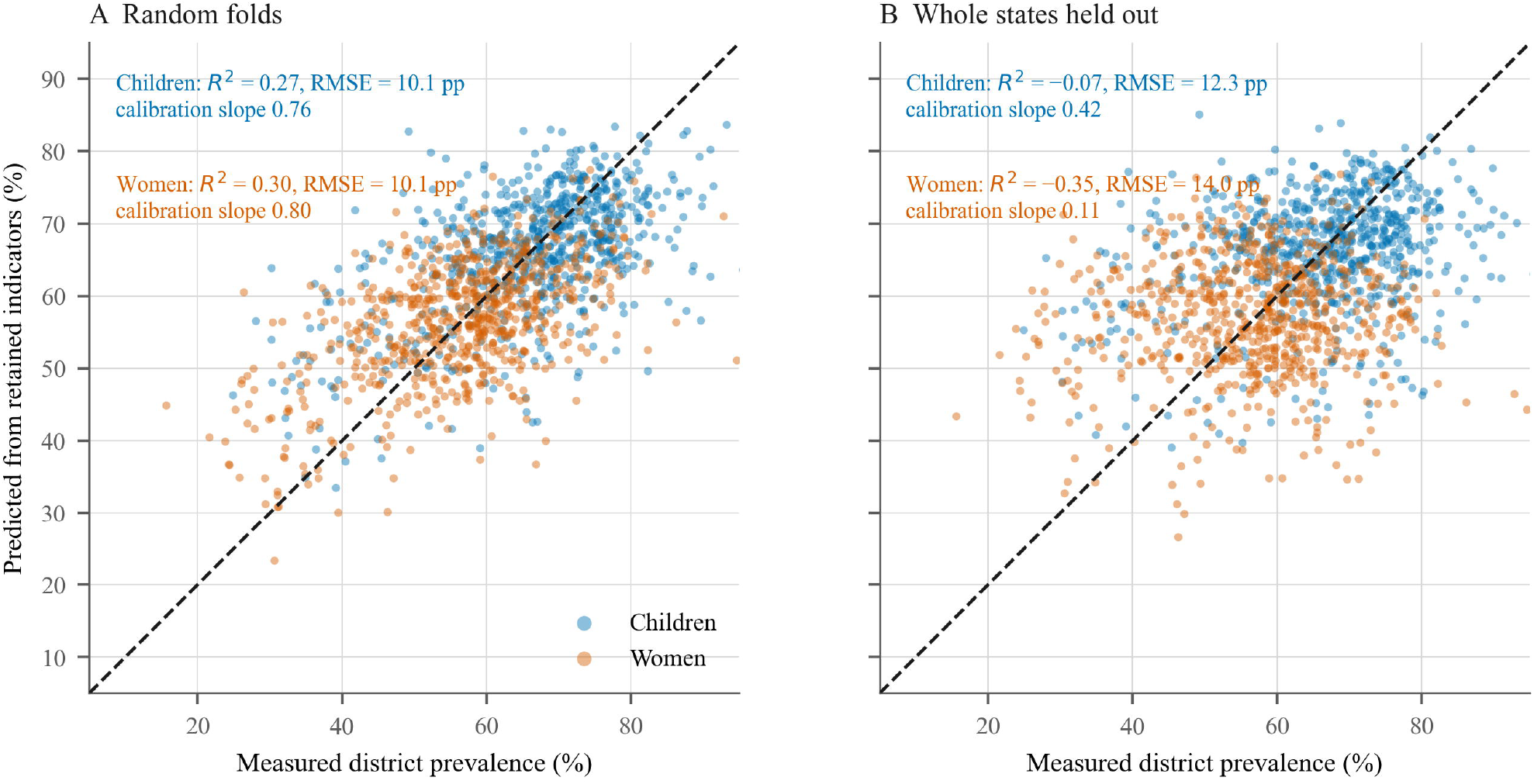
Predicting district anaemia from the indicators NFHS-6 retained. Each point is one district; the dashed line is equality. Predictions come from gradient-boosted regression trees using only the indicators the NFHS-6 fact sheets still report, and are out of sample in both panels. (A) Ten-fold cross-validation over districts. (B) Whole States held out, so no district of the target State is seen in training. Both panels annotate both populations: children in the upper label, non-pregnant women in the lower. R^2^ is the out-of-sample coefficient of determination and RMSE the root mean squared error in percentage points; a negative R^2^ means the surrogate predicts worse than the training-fold mean. R^2^ = 0.27, women R^2^ = 0.30. Held-out States: children R^2^ = −0.069, women R^2^ = −0.355. Calibration slopes appear in each panel; a slope below one means predictions are compressed towards the mean.

Within States, only one input was associated with lower anaemia once seven were tested: women’s schooling, at −1.67 pp per 10 pp for children (95% confidence interval −2.57 to −0.78) and −1.18 pp for women (−1.99 to −0.38). It is a structural characteristic, not a programme service, and no service indicator reached that standard; vitamin A in women was closest, at −0.98 pp (−1.72 to −0.25). Applying the associations to the national gains, through the covariance of the coefficients and the three indicators reported in both rounds, implied a change in child anaemia of +0.01 pp (−0.91 to +0.92) and in women of +0.50 pp (−0.33 to +1.33).

### Step 3. What changes in prioritisation

Under the full NFHS-5 set, 10 districts were robust priorities for children; under the reduced set, 13. Membership shifted more than the totals let on: 49 of 707 districts changed class and 15 of 71 top-decile choices differed (Fig. 2). For women, 62 districts changed class and 19 choices differed. Removing anaemia alone still moved 32 districts and 10 child choices.

**Figure 2.**
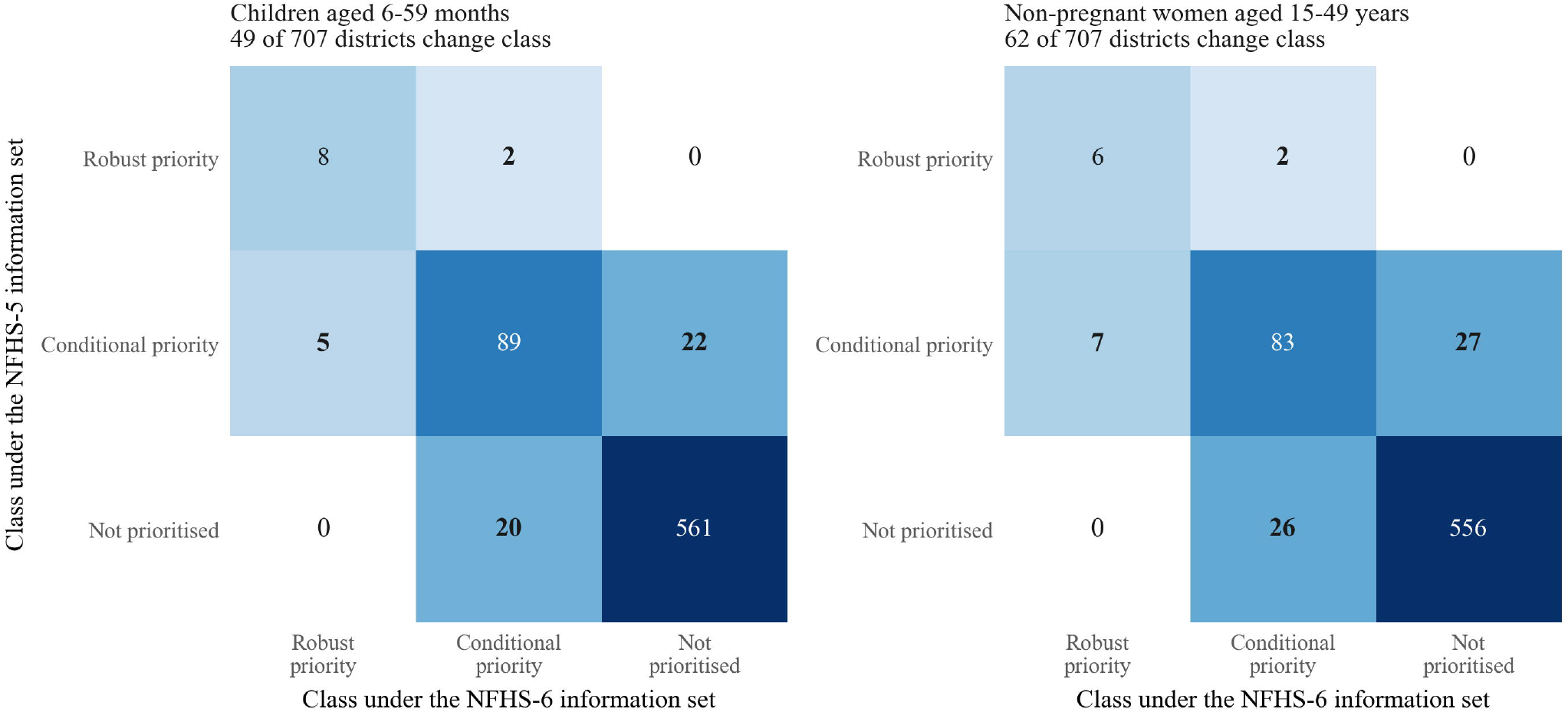
Movement of districts between priority classes when the discontinued indicators are removed. Rows are the priority class under the full NFHS-5 information set, columns the class under the indicators NFHS-6 retains; cells give the number of districts, and shading is proportional to the logarithm of that count. The diagonal holds districts whose classification survives the loss; every off-diagonal cell is a district the programme would treat differently. 49 of 707 districts change class for children and 62 for women. Priority classes come from stochastic multicriteria acceptability analysis over 10 000 draws of the criterion posteriors and of a hierarchical weight vector, with the same preference draws used under both information sets.

More revealing is how far the composite can see its own uncertainty. Under the full information set, 3.4 of 71 selections for children lay outside the top set implied by the drawn criterion values. Under the reduced set the figure is zero — not because the choice improved, but because anaemia was the only criterion whose uncertainty this analysis could propagate. With it gone, the composite returns a perfectly confident ranking, a pattern unchanged by the class boundaries or the preference model.

### Step 4. What the uncertainty costs

Of 71 districts a programme would select for intensive child anaemia action on current estimates, 19.1 fall outside the latent top 71 — 26.9% — and knowing the latent values would raise the chosen set’s mean burden by 1.02 pp. For women, 8.4 of 71, or 11.8%. Varying the decision from 5% to 20% of districts left this between 22.4% and 26.9% for children.

Which districts are measured matters more than how many (Fig. 3). At 183 districts, the four rules using the decision recovered 46.1% to 46.3% of the gap for children and 44.6% for women, with replicate ranges under a percentage point. The widest posteriors recovered 22.0% and 22.7%: being unsure about a district is related to, but not the same as, needing to know. The four frame-based allocations recovered 9.6% to 13.3% for children, with ranges from 6.4% to 17.2% overlapping too heavily to support any ordering among them. A well-chosen network of 100 districts already recovered 43.3%, with returns flattening beyond about 150 — conditional on this ordering, precision and decision.

**Figure 3.**
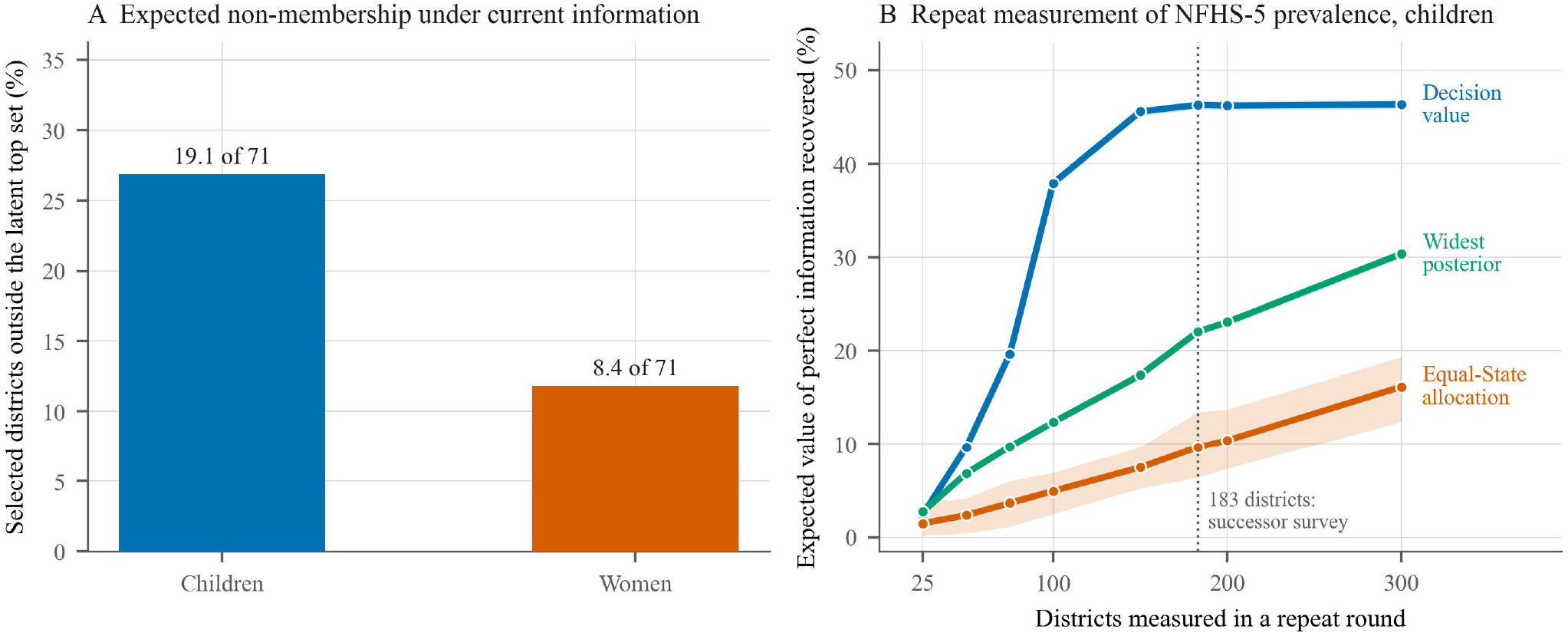
Decision uncertainty in district targeting, and how the choice of measured districts changes it. (A) Expected non-membership: of the 71 districts a programme would select on posterior mean prevalence, the number and percentage falling outside the latent top 71 in a posterior draw — 19.1 districts (27%) for children and 8.4 (12%) for women. This is a property of the posterior, not a count of observed errors. (B) Share of the expected value of perfect information recovered by a repeat measurement of each network size, for children, under one rule that uses the decision and two that do not. Curves are labelled at their right-hand ends: *Decision value* selects districts by their per-district expected value of partial perfect information; *Widest posterior* selects those whose posterior standard deviation is largest; *Equal-State allocation* takes one district from each State or Union Territory in turn. Shaded bands are the 2.5th to 97.5th percentile across 40 replicates, in which rules that need a frame draw a new one and every rule draws new survey outcomes. Districts are chosen using one half of the posterior draws and the network scored on the other. Each measured district is observed subject to its own design-based sampling error, on the logit scale. The dotted line marks 183 districts, the number the successor survey covers. The calculation values a repeat measurement of the 2019-21 latent prevalence under stationarity and an unchanged measurement platform; it does not evaluate that survey’s own district list, which is not public.

Two things change that appraisal. Allowing district values to have moved since 2019–21 raises what measurement recovers, to 63.9% at a movement of 5 pp, because a stale estimate is then wrong twice over. A platform difference does the opposite: a venous reading 10 pp lower than the capillary-anchored prior, varying by district, cuts recovery to 33.0% for children and 9.8% for women, and leaves the frame-based rules worse than not measuring. Halving or doubling the sample per district moved recovery between 30.8% and 62.3%, so achieved precision matters about as much as the rule.

Applying current age-specific child cut-offs instead of the survey’s single threshold lowered the median district by 3.33 pp but left the ordering nearly intact, at a rank correlation of 0.991 with 65 of 71 top-decile districts unchanged.

## Discussion

Three findings follow. Programme input coverage cannot reproduce measured district anaemia: a flexible model given every surviving indicator carries a structural error two and a half to four times the sampling error of measurement, compresses its predictions towards the mean, and in an unseen State does worse than its own training mean. Losing the indicator moves which districts a composite selects and, more importantly, removes the composite’s only means of reporting how uncertain that selection is. And where measurement must be bounded, the choice of districts dominates the count: at the size now fielded, choosing for effect on the decision recovered about four times as much as any allocation built around coverage of States.

The limits matter, because the most quotable numbers are the most conditional. Step 4 conditions on the NFHS-5 posterior and values a repeat measurement of that same latent prevalence, on the same platform, under stationarity. It is not an estimate of what removing anaemia from NFHS-6 has cost, contains no model of change since 2019–21, and does not evaluate the successor survey’s district list, frame or achieved precision, none of which is public. The split-sample check is not external validation, since both halves come from one posterior. Step 2 was chosen after the data had been seen and should be treated as a finding to be replicated. Uncertainty was propagated only for anaemia, since design-based variances existed only there, which makes the priority classes look steadier than they are. The analysis is ecological and supports no causal claim; district coverage is a weak proxy for dose, duration and adherence, and supplementation follows need, so confounding by indication is likely. District denominators were unavailable, so priority reflects prevalence rather than people affected, and the two rounds use different district frames, 707 against 715. Against these limits, the study covers every district, derives precision from the microdata, tests the surrogate against unseen States, and regenerates every quantity from code.

That service indicators fail to reproduce a haemoglobin-based outcome is consistent with wider evidence that haemoglobin alone identifies neither iron deficiency nor its alternatives, and that the biomarkers needed to direct treatment are not collected by routine reporting.^19,20^ Our result is narrower: the indicators are not uninformative, but their error is several times that of measuring the outcome.

The findings generalise to district-level targeting in India under the measurement model stated, and no further; the held-out-State analysis speaks to transfer across regions, the scenarios to transfer across platforms and time. Three implications follow. A bounded venous network is where a second selection criterion is cheapest to add: sorting candidate districts additionally by distance from an action threshold would serve targeting without displacing the estimation the survey exists for, though changing which districts enter has consequences for representativeness, precision and field cost. Outside such a network, input coverage must not be read as an outcome; the last measured values remain the best basis for planning, but with credible intervals about twelve percentage points wide they should mark out priority classes, not league tables. And the Track pillar needs a population outcome, not only a service record, since a portal pooling haemoglobin from clinical platforms captures the people who turn up, who are not a fair sample.

## Conclusion

India has improved how it measures anaemia and reduced where. The gain in accuracy is real. What this analysis shows is that the surviving programme indicators cannot rebuild district anaemia — their structural error is several times that of measurement, and they do not travel to unseen States — and that rankings built on the last available measurement are uncertain enough that more than a quarter of a selected set would not belong in it.

Rules choosing districts for their effect on the decision recover several times more of that uncertainty than any allocation built around spreading a sample across States, while precision and platform comparability matter as much as the rule. A programme organised around tracking should treat the make-up of its measurement network as a decision in its own right, and establish that a new platform is comparable before using it to rank districts.

## Supporting information

STROBE reporting guidelines

## Declarations

### Ethics approval

Not applicable. This is a secondary analysis of published survey fact sheets and de-identified microdata; no human participant was contacted or recruited, and no identifiable information was accessed at any stage. Under the Indian Council of Medical Research National Ethical Guidelines for Biomedical and Health Research Involving Human Participants (2017) and the institution’s own policy, secondary analysis of de-identified data does not require Ethics Committee review. None was therefore sought.

### Consent

Informed consent, including consent for anaemia testing, was obtained from every participant by the original survey under its own protocol. De-identified individual-level records were used here only to compute sampling variances, and no individual is identifiable in any output. No additional consent was sought for this secondary analysis, which the data-access terms permit.

### Author contribution

The sole author conceived the study, obtained and validated the data, designed and implemented the analysis, produced the figures and tables, wrote the manuscript, and accepts full responsibility for the integrity of the data and the accuracy of the analysis.

### Prior publication

The manuscript has not been published elsewhere, in whole or in part, and is not under consideration by any other journal.

### Data quality note

The widely circulated compilation of the NFHS-5 district fact sheet contains 707 rows but not 707 distinct districts: Manipur’s Chandel appears twice, once relabelled as a district of Mizoram with all 107 indicator values duplicated, while Mizoram’s Saiha is absent. Analyses built on that file without checking will carry one State’s values into another. The row was relabelled and that district’s criteria left missing, which the model treats as genuine ignorance.

### Data availability

The NFHS-6 fact sheets and the NFHS-5 district fact sheet are published by the International Institute for Population Sciences and are freely available. The NFHS-5 person recode is not public: it is distributed by the DHS Program under registered controlled access, and a registered user can obtain the identical file. Because redistribution is not permitted, the derived district-level quantities extracted from it are provided instead, which is sufficient to reproduce every result reported here. The analysis code, the derived data and a checksum manifest accompany this submission and will be deposited in a public repository with a persistent identifier on acceptance.

