## Supplementary file for "Targeting anaemia without measuring it: surrogate prediction, district decision uncertainty and the value of repeat measurement in India"

This material carries the detail the word limit kept out of the main text. It is generated directly from the analysis output, so every number here matches the manuscript by construction. Sections S1 to S4 follow the four analytic steps in the same order; S5 to S9 cover the diagnostic-threshold sensitivity analysis, data provenance, a model that was planned and abandoned, and reproduction.

### S1. How district anaemia and its uncertainty were estimated

#### S1.1 Why a model was needed at all

The median district contributed 243 measured children and 941 measured women. A proportion from a few hundred people, collected in clusters rather than at random, is imprecise. Treating such a figure as though it were known exactly is what produces unstable district league tables, in which a district's rank changes from one round to the next for reasons that are purely statistical.

#### S1.2 Step-by-step procedure

1. **Extract the raw measurements.** From the NFHS-5 person recode, take every

person with a valid haemoglobin reading who slept in the household the previous night, which is the population the survey itself tabulates. This gives 1 685 994 person-records.

2. **Define anaemia.** Children aged 6–59 months with haemoglobin below

11.0 g/dl; non-pregnant women aged 15–49 years below 12.0 g/dl. These are the survey's own operational definitions, chosen because the estimand is the published district figure a programme works from. Section S5 reports what happens under the age-specific cut-offs of the 2024 WHO guideline. Values are the altitude-adjusted ones the survey supplies.

3. **Compute the design-based standard error for each district.** Apply the

survey's sampling weights, strata and primary sampling units, and use Taylor linearisation. This accounts for the fact that people in the same cluster resemble one another, which a simple binomial calculation ignores.

4. **Check the extraction against the published values.** For children the two

routes correlated 0.98997 with a mean absolute difference of 1.158 pp, a mean signed difference of 0.469 pp and limits of agreement -2.903 to 3.841 pp; 82.9% of districts agreed within 2 pp. For women the correlation was 0.99995, the mean absolute difference 0.089 pp and the mean signed difference 0.031 pp. The extraction also reproduced the survey's own anaemia classification variable exactly. Agreement is reported rather than correlation alone, because a high correlation is compatible with a systematic offset.

5. **Transform to the logit scale.** Prevalence is bounded between 0 and 1;

modelling it on the logit scale prevents credible intervals running past those bounds. The standard error is carried across by the delta method.

6. **Fit the Fay–Herriot model** with districts nested in States, the

observation variance fixed at the design-based value from step 3, and half-Cauchy priors on the variance components.

7. **Sample.** Four Gibbs chains, 1000 burn-in iterations discarded, 4000 draws

retained per chain, giving 16 000 draws per district.

#### S1.3 Convergence, efficiency and predictive checks

The largest potential scale reduction factor across all 707 district effects was 1.0005 for children and 1.0005 for women. Because a scale reduction factor near one can coexist with slow mixing, effective sample sizes are given for every hyperparameter.

| **Parameter** | **Children: R-hat** | **Children: effective n** | **Women: R-hat** | **Women: effective n** |
| --- | --- | --- | --- | --- |
| District variance τ² | 1.00013 | 5613 | 1.00004 | 9059 |
| State variance ω² | 1.00005 | 5033 | 1.00007 | 4174 |
| Grand mean μ | 1.00574 | 120 | 1.00887 | 72 |

The variance components mix well. The grand mean is the slowest-mixing parameter, with an effective sample size of 120 for children and 72 for women out of 16 000 nominal draws, which is what a flat prior on a level parameter shared by 707 nested units produces. Nothing reported in this paper depends on the level: every downstream quantity is a rank, a difference or a set membership, all of which are invariant to a common shift.

A posterior predictive check replicated a full set of district estimates from the fitted district effects and the known design variances at every iteration, and compared their spread with the spread of the real estimates. The replicated standard deviation on the logit scale averaged 0.5489 against an observed 0.5658 for children, giving a tail probability of 0.0487; for women the figures were 0.5201 against 0.5244, tail probability 0.2491. The child model is therefore at the edge of under-dispersion: it reproduces slightly less spread among districts than the data show, which would make the child posteriors marginally too smooth and, if anything, understate how far apart districts are.

Refitting under half-Cauchy prior scales of 0.5 and 2.0 moved no district posterior mean by more than 0.343 pp, with a mean absolute shift of 0.0667 pp. The posterior is not a product of its prior scale.

#### S1.4 Singleton strata

A stratum contributing a single primary sampling unit within a district supplies no between-cluster contrast. The analysis drops those contributions, which understates the variance of the districts affected. 16 districts (2.3%) contain at least one such stratum for children. Recomputing their variance with the lone unit centred on the district's overall contribution — the standard adjustment — raises their median standard error from 4.043 pp to 4.102 pp, a median ratio of 1.003. The understatement is therefore real but small, and it works against this paper's argument, since a wider sampling variance would make measurement more valuable rather than less.

#### S1.5 What the model does to an estimate

A district measured on few children is pulled towards its State average; a well-measured district is left where it is. Averaged over all districts, the standard deviation fell from 3.662 pp to 3.283 pp for children and from 2.304 pp to 2.178 pp for women. The median 95% credible interval remained 12.62 pp wide for children and 8.29 pp for women, which is the honest width of what is known about a district.

### S2. Testing whether the surviving indicators can substitute

#### S2.1 The question, stated as a test

If a statistical model could reproduce district anaemia from the indicators NFHS-6 still reports, then discontinuing the measurement would cost little: the number could be inferred instead of measured. This is a prediction problem, and it has a right way to be judged — out of sample, on districts the model has not seen.

#### S2.2 Step-by-step procedure

1. **Assemble the predictor sets.** Set A holds the seven prioritization criteria NFHS-6 retains. Set B adds every remaining NFHS-5 indicator, including sanitation, cooking fuel, stunting, underweight, institutional delivery and health insurance, so that a poor result cannot be blamed on withholding information.

2. **Fit three model classes.** A mean-only baseline, refitted inside each training fold so that it too is scored out of sample; a penalised linear model with the penalty chosen by internal cross-validation; and gradient-boosted regression trees, which can capture interactions and non-linearity.

3. **Validate two ways.** Random ten-fold cross-validation, and grouped cross-validation that withholds every district of a State at once.

4. **Handle missing predictors by complete case.** Of 707

districts, 14 lack at least one predictor and are excluded, leaving 693. The exclusions are dominated by a single indicator: full immunisation is missing for 14 districts. One of those is the repaired fact-sheet row described in S6.3, whose values were cleared because they belonged to another State.

#### S2.3 Comparing prediction error with measurement error

The prediction is scored against published district estimates, which are themselves measured with error. The raw discrepancy therefore contains both the model's error and the survey's, and dividing it by a median standard error would compare a root-mean-square quantity across heterogeneous districts with a within-district median. Both problems are corrected here.

The mean squared error of the outcome's own sampling variation is subtracted from the observed mean squared error, leaving the structural component attributable to the model, and that component is compared with the root-mean-square district standard error. For children the root mean squared error was 10.14 pp and the mean absolute error 7.85 pp; after removing the outcome's sampling variance the structural error was 9.44 pp against a root-mean-square district standard error of 3.72 pp, a ratio of 2.5. For women the corresponding figures were 10.08 pp, 7.96 pp, 9.80 pp, 2.34 pp and 4.2.

District standard errors are not interchangeable with their median. For children they ran from 1.209 pp to 7.223 pp, with quartiles 3.012 and 4.123 pp around a median of 3.537 pp and a root-mean-square value of 3.721 pp.

#### S2.4 Why the two validation schemes differ

Random folds leave a model with neighbouring districts of the same State in its training data, so it can infer the State's level and interpolate. Grouped folds remove that crutch. Because between-State variation dominates district anaemia in India, the grouped result is the one that describes a surrogate asked to stand in for a discontinued national measurement — a situation in which whole regions may be unmeasured.

#### S2.5 Associations between inputs and anaemia, within States

Estimated with State fixed effects and heteroskedasticity-consistent standard errors, for both populations. A positive value means higher coverage accompanies *more* anaemia. Seven indicators are tested at once, so a Holm-adjusted p value is given. These are ecological associations, not effects, and no causal reading is available from them.

| **Population** | **Indicator** | **Change in anaemia (pp) per 10 pp coverage** | **95% CI** | **p (Holm)** |
| --- | --- | --- | --- | --- |
| Children | anc4 | 0.472 | -0.217 to 1.161 | 0.71619 |
| Children | ifa100 | -0.37 | -1.135 to 0.395 | 0.87552 |
| Children | full immun | 0.083 | -0.669 to 0.835 | 0.87552 |
| Children | vitamin a | -0.711 | -1.558 to 0.136 | 0.49895 |
| Children | adequate diet | 0.65 | -0.559 to 1.859 | 0.87552 |
| Children | menstrual hygiene | -0.82 | -1.72 to 0.079 | 0.44269 |
| Children | woman schooling | -1.673 | -2.568 to -0.778 | 0.00175 |
| Women | anc4 | 0.384 | -0.213 to 0.981 | 0.8285 |
| Women | ifa100 | 0.031 | -0.635 to 0.697 | 1 |
| Women | full immun | 0.198 | -0.475 to 0.87 | 1 |
| Women | vitamin a | -0.983 | -1.716 to -0.251 | 0.05112 |
| Women | adequate diet | -0.338 | -1.365 to 0.689 | 1 |
| Women | menstrual hygiene | -0.699 | -1.539 to 0.141 | 0.51368 |
| Women | woman schooling | -1.184 | -1.992 to -0.376 | 0.02845 |

#### S2.6 The national projection, and what it is not

Applying these cross-sectional coefficients to the national gains in coverage between rounds gives +0.01 pp for children (95% CI -0.91 to +0.92) and +0.50 pp for women (-0.33 to +1.33). The interval is computed from the full robust covariance matrix of the coefficients, because they are correlated and combining their marginal intervals as if independent would misstate the interval on their sum.

Only 3 of the seven retained indicators carry a published value in both rounds (anc4, ifa100, full_immun), so the combination is partial as well as cross-sectional. It is a descriptive statement about what the observed associations would imply if they held over time. They are not known to hold over time, and the quantity is not a forecast of prevalence, is not offered as one, and appears in the main text only because it is indistinguishable from zero.

### S3. Prioritisation under three information sets

#### S3.1 How weights were handled

Any composite score depends on how its criteria are weighted, and any particular set of weights is a value judgement a reader may reject. Stochastic multicriteria acceptability analysis samples the weight space instead of fixing it. That does not amount to choosing no weights: a weight distribution is still a model, and the one used here is stated rather than implied.

Weights are drawn hierarchically. A share is drawn for each of the four domains from a flat Dirichlet, then split within the domain among its own criteria. Drawing ten criterion weights flat from a simplex would give the delivery domain four times the expected weight of the burden domain purely because it contains four indicators, which is an accident of bookkeeping rather than a preference anyone holds.

Comparisons across information sets use matched preference draws. The same underlying domain and within-domain proportions are used in every world; when a criterion is unavailable its weight is removed and what remains is renormalised. Any difference between worlds is then attributable to the missing information and not to a differently shaped preference distribution.

#### S3.2 Step-by-step procedure

1. **Orient every criterion** so that a higher value means a greater claim on

attention. Coverage indicators are therefore entered as deficits: a district with 40% antenatal coverage enters as 60.

2. **Draw criterion values.** Anaemia comes from the S1 posterior draws; the

others are taken at their published values. Missing values are drawn from the district's own State, so a district with no data widens rather than disappears.

3. **Draw a hierarchical weight vector** as described above, from a fixed stream

independent of the value draws.

4. **Normalise and rank** by TOPSIS, which scores each district by its distance

from the best and worst attainable profiles.

5. **Record** whether the district reached the top tenth.

6. **Repeat 10 000 times** and take the proportion as the district's

acceptability.

7. **Classify.** Robust priority at 80% or more; conditional between 20% and

80%; otherwise not prioritised.

8. **Repeat the whole procedure** under each of the three information sets, with

matched preference draws, and cross-tabulate the classifications.

#### S3.3 What changed, in classes and in the underlying score

For children, 49 of 707 districts changed class between the full and reduced information sets, and 15 of 71 top-decile selections differed. Removing anaemia alone moved 32 districts and 10 selections.

Class counts can hide movement, so the continuous score is reported too. The mean absolute change in acceptability between the full and reduced sets was 0.0428 for children, and the 90th percentile of that change was 0.1483.

#### S3.4 Sensitivity to the class boundaries (children)

| **Class boundaries** | **Robust priorities, NFHS-5 set** | **Robust priorities, NFHS-6 set** | **Districts changing class** |
| --- | --- | --- | --- |
| 0.10 / 0.90 | 1 | 6 | 62 |
| 0.20 / 0.80 | 10 | 13 | 49 |
| 0.30 / 0.70 | 22 | 22 | 57 |

#### S3.5 Sensitivity to the model of preferences (children)

| **Preference model** | **Robust priorities, NFHS-5 set** | **Robust priorities, NFHS-6 set** | **Districts changing class** | **Top-decile selections differing** |
| --- | --- | --- | --- | --- |
| concentrated preferences (Dirichlet 0.5 across domains) | 1 | 9 | 53 | 15 |
| evenly spread preferences (Dirichlet 4 across domains) | 32 | 34 | 55 | 16 |
| equal weight to each domain | 32 | 28 | 50 | 15 |
| half the weight on burden | 29 | 22 | 114 | 38 |

The reference model is a flat Dirichlet across domains. Concentrating or spreading preferences changes little. Fixing half the weight on the burden domain changes a great deal, which is the expected and correct behaviour: the more a decision maker cares about the outcome, the more it matters that the outcome is no longer observed.

#### S3.6 A decision that stops being able to see its own uncertainty

Within each iteration the same weights were applied twice: once to the drawn criterion values, treated as the underlying truth, and once to the posterior means, which is what a programme would act on. The number of selected districts falling outside the drawn top set is the decision cost of the remaining uncertainty, measured under the multicriteria rule rather than on burden alone.

Under the full NFHS-5 information set this was 3.44 of 71 for children. Under the reduced set it is 0.0.

That zero is not an improvement and should not be read as one. Design-based variances were available only for the haemoglobin-based indicators, so anaemia is the only criterion whose uncertainty this analysis propagates. Remove it and the composite has no uncertainty left to propagate, and returns a perfectly confident ranking. The apparent certainty is a property of the arithmetic, not of the evidence, and it is the clearest available illustration of what is lost when the measured outcome leaves a scoring system: not the score, but the score's ability to report its own unreliability.

### S4. The decision value of measurement, and where it would be spent

#### S4.1 What is and is not estimated

Value of information has no meaning without a decision. The decision here is: select 71 districts, one tenth, for intensive action, with a district's benefit taken as proportional to its true anaemia prevalence.

The estimand is expected non-membership under the NFHS-5 posterior: of the 71 districts a programme would select on posterior mean prevalence, how many fall outside the latent top 71 in a posterior draw. The measurement being valued is a repeat observation of that same 2019–21 latent prevalence, on the same measurement platform, with each district's own design-based sampling error.

Four things are therefore **not** estimated anywhere in this paper. There is no model of how district prevalence changed between 2019–21 and the present. There is no capillary-to-venous bridge in the reference calculation. There is no use of the successor survey's district list, sampling frame or achieved precision, none of which is public. And there is no counterfactual round in which anaemia had been retained. No quantity here is the realised cost of any decision taken by any agency. Section S4.5 relaxes the first two assumptions explicitly.

#### S4.2 Step-by-step procedure

1. **Split the posterior draws in half.** Everything used to *choose* districts

comes from the design half, including the per-district decision value that orders the leading rule; everything used to *score* the resulting network comes from the evaluation half. This checks that a rule is not flattered by the simulation that produced it. It is not an external validation: both halves are draws from the same fitted posterior, so it says nothing about how the rule would perform against new data or a later survey.

2. **Compute the current decision.** Rank districts by posterior mean and take

the top 71.

3. **Compute the perfect decision.** Within each draw, take the latent top 71.

4. **Measure the gap** as the expected number of selected districts outside that

latent set, and as the expected value of perfect information.

5. **Score eight selection rules** at network sizes from 25 to 300 districts.

6. **Model measurement on the scale the model is fitted on.** A measured

district returns an estimate, not the truth. The estimate is generated and combined with the prior on the logit scale by precision weighting, with each district carrying its own design-based variance rather than a shared median.

7. **Replicate.** Every rule is scored 40 times.

Rules that need a frame draw a new one each time; every rule draws new survey outcomes. The range across replicates is reported.

8. **Vary the decision size** from 5% to 20% of districts, and vary the

measurement and stationarity assumptions (S4.5).

#### S4.3 All rules at the size the successor survey covers (183 districts)

Children:

| **Selection rule** | **Recovered (%)** | **95% range across replicates** |
| --- | --- | --- |
| decision value (EVPPI) | 46.3 | 45.8 to 46.7 |
| highest estimated burden | 46.2 | 45.7 to 46.6 |
| closest to the selection boundary | 46.1 | 45.7 to 46.4 |
| widest posterior | 22 | 21.6 to 22.4 |
| allocation proportional to measured population | 13.3 | 9.0 to 17.2 |
| simple random | 13.2 | 10.0 to 16.5 |
| allocation proportional to district count | 12.9 | 10.1 to 16.6 |
| equal allocation across States and UTs | 9.6 | 6.4 to 13.5 |

Non-pregnant women:

| **Selection rule** | **Recovered (%)** | **95% range across replicates** |
| --- | --- | --- |
| decision value (EVPPI) | 44.6 | 44.1 to 45.2 |
| closest to the selection boundary | 44.6 | 43.9 to 45.5 |
| highest estimated burden | 44.6 | 43.9 to 45.3 |
| widest posterior | 22.7 | 22.0 to 23.2 |
| allocation proportional to district count | 11.3 | 5.7 to 17.1 |
| allocation proportional to measured population | 11.3 | 6.5 to 16.6 |
| simple random | 10.7 | 4.8 to 17.3 |
| equal allocation across States and UTs | 10.2 | 4.7 to 16.0 |

Three groups appear. Rules that use the decision recover about the same amount as each other, with narrow ranges. Choosing the districts whose posterior is widest recovers roughly half as much: being unsure about a district is related to, but not the same as, needing to know. The four frame-based allocations are indistinguishable from one another, and their ranges overlap so heavily that no ordering among them is supported.

The reported curves are essentially monotone in network size; residual movement of about 0.1 percentage points at the largest sizes is Monte Carlo noise and is covered by the reported ranges.

#### S4.4 Sensitivity to the size of the decision (children)

| **Districts selected** | **Expected non-membership (n)** | **Expected non-membership (%)** |
| --- | --- | --- |
| 35 | 9.02 | 25.8 |
| 71 | 19.09 | 26.9 |
| 141 | 31.59 | 22.4 |

#### S4.5 Sensitivity to the measurement and stationarity assumptions

Each row relaxes one assumption of the reference calculation, at 183 districts. Platform bias is the shift a venous reading would show against the capillary-anchored prior, expressed in percentage points at median prevalence. Drift is independent district movement since the round the prior describes. The sample multiplier scales the achieved sample per district.

Children:

| **Scenario** | **Decision-led rule (%)** | **Equal-State allocation (%)** |
| --- | --- | --- |
| same platform, no drift (reference) | 46.3 | 9.7 |
| venous reads 5 pp lower than capillary | 46.1 | 4.1 |
| venous reads 10 pp lower, varying by district | 33 | -6.5 |
| district values have moved (SD 5 pp) | 63.9 | 15.4 |
| district values have moved (SD 10 pp) | 69.7 | 21.9 |
| half the sample per district | 30.8 | 6.2 |
| twice the sample per district | 62.3 | 13.4 |
| moved 5 pp and venous reads 5 pp lower | 63.6 | 10.5 |

Non-pregnant women:

| **Scenario** | **Decision-led rule (%)** | **Equal-State allocation (%)** |
| --- | --- | --- |
| same platform, no drift (reference) | 44.7 | 10.2 |
| venous reads 5 pp lower than capillary | 44.7 | -4.3 |
| venous reads 10 pp lower, varying by district | 9.8 | -49.4 |
| district values have moved (SD 5 pp) | 65.5 | 16.2 |
| district values have moved (SD 10 pp) | 70.5 | 20.1 |
| half the sample per district | 28.2 | 6.2 |
| twice the sample per district | 62.1 | 14.3 |
| moved 5 pp and venous reads 5 pp lower | 65.4 | 11.7 |

Three readings follow. Measurement is worth more, not less, once district values are allowed to have moved, because a stale estimate is then wrong for two reasons rather than one. Achieved precision matters roughly as much as the choice of rule at these sample sizes. And an uncorrected platform difference that varies by district can destroy the value of the exercise altogether, and in the worst row leaves the decision worse than not measuring: measured districts are then shifted relative to unmeasured ones on a scale that has nothing to do with their burden. Any new platform therefore needs a bridging study before its readings are used to rank districts against districts measured the old way.

### S5. Does the diagnostic threshold change the answer?

The main analysis uses the survey's operational definition, a single cut-off of 11.0 g/dl for every child aged 6–59 months. The 2024 WHO guideline instead sets 10.5 g/dl below 24 months and 11.0 g/dl from 24 months. Age in months was re-extracted from the person recode for all 184,184 measured children, of whom 32.2% were under 24 months, and district prevalence recomputed under both rules with the same variance estimator.

| **Quantity** | **Value** |
| --- | --- |
| Median district prevalence, operational definition | 68.47% |
| Median district prevalence, WHO 2024 age-specific | 64.84% |
| Median difference | -3.33 pp |
| Spearman rank correlation between definitions | 0.9906 |
| Top-decile districts shared | 65 of 71 |

The level falls, as it must, since the younger cut-off is lower. The ordering barely moves. Because every quantity in this paper is a rank, a class or a set membership rather than a level, the choice of operational definition is not what drives the findings. The main results retain the survey's own definition because the estimand is the published figure a programme works from, not a re-estimate of prevalence.

### S6. Data provenance and a defect in a widely used file

#### S6.1 Sources

| **Source** | **Used for** | **Level** |
| --- | --- | --- |
| NFHS-5 district fact sheet, 2019-21 | Anaemia prevalence; ten prioritisation criteria | District |
| NFHS-5 person recode | Design-based standard errors, design effects, sample sizes, age in months | Individual |
| NFHS-6 fact sheets, 2023-24 | Which indicators survive into the current round | State and national |

#### S6.2 What the sixth round does and does not report

An automated text probe of all 182 pages of the released fact sheets found the following. Present: adequate diet, antenatal, blood sugar, full immunisation, institutional birth, iron folic acid, menstrual, schooling, stunted, tobacco, underweight, vitamin a, wasted. Absent: anaemia, clean fuel, haemoglobin, sanitation. The document was checksummed on download and the SHA-256 digest is recorded with the analysis output.

#### S6.3 A duplicated district in the circulating fact-sheet compilation

The widely circulated compilation contains 707 rows but not 707 distinct districts. Manipur's Chandel appears twice, once relabelled as a district of Mizoram, with all 107 of 107 indicator values identical. Mizoram's Saiha is absent. Any analysis using that file without checking will carry one State's values into another. The row was relabelled to Saiha and its indicator values cleared, so the district enters the analysis with missing criteria that the model treats as genuine ignorance rather than as an average.

### S7. A model that was planned and abandoned

The analysis protocol specified an efficiency analysis: districts as units converting service contact into anaemia-free children, estimated by a stochastic frontier with inefficiency effects and cross-checked by data envelopment analysis. It is recorded here rather than dropped, so that a reader comparing the protocol with the paper can find it.

| **Population** | **γ** | **σ_v** | **Elasticities with the wrong sign** | **Identified** |
| --- | --- | --- | --- | --- |
| Children | 1.0 | 2.9e-05 | 3 of 5 | No |
| Women | 1.0 | 2.4e-05 | 4 of 5 | No |

γ is the share of residual variance attributed to inefficiency. A value of 1.000 with σ_v at essentially zero means the likelihood has assigned the entire residual to inefficiency and none to noise. Combined with wrong-signed elasticities, this shows the model does not fit. No district is described as efficient or inefficient anywhere in this work.

What this result does *not* do is explain itself. Non-identification is consistent with inputs that do not track the outcome, with a misspecified functional form, with measurement error in the inputs, and with several other causes that point in different directions. It is therefore recorded for completeness and carries no weight in the argument of this paper.

The substitutability analysis in S2 replaced it. That analysis was chosen after the data had been seen and is exploratory rather than confirmatory, which is why it was judged by the more demanding held-out State standard.

### S8. Reproducing this analysis

#### S8.1 What is needed

- Python 3.14 with numpy, pandas, scipy, scikit-learn, matplotlib, pyarrow and

python-docx.

- The NFHS-5 person recode, obtainable by any registered user from the DHS

Program under its data-access terms. Redistribution is not permitted, so it is not included; the derived district-level quantities extracted from it are.

- The NFHS-5 district fact sheet and the NFHS-6 fact sheets, both public.

#### S8.2 Order of execution

| **Step** | **What it does** |
| --- | --- |
| 1 | Link microdata district codes to named districts and validate the linkage |
| 2 | Extract child haemoglobin with age in months for the threshold analysis |
| 3 | Download the NFHS-6 fact sheets and verify which indicators they report |
| 4 | Assemble the analytic district dataset |
| 5 | Run the four analytic steps and the threshold sensitivity analysis |
| 6 | Build tables and figures |
| 7 | Number references by order of first mention |
| 8 | Check every manuscript number against the analysis output |
| 9 | Generate the submission package |

The pipeline is deterministic under a fixed seed (20260730).

#### S8.3 Verification built into the pipeline

Step 8 re-reads the analysis output and confirms that every numerical claim in the manuscript still matches it, that the journal's word and display limits are met, that references are numbered in order of first mention and cited in the journal's style, and that no placeholder text remains. A failure stops the build.

### S9. Supplementary tables provided as separate files

| **File** | **Contents** |
| --- | --- |
| supp_table_district_results.csv | Every district: prevalence, standard error, sample size, acceptability and class under each information set, and per-district decision value |
| supp_table_associations.csv | Within-State associations between each input and anaemia, both populations, with Holm-adjusted p values |
| supp_table_sentinel_designs.csv | All eight selection rules at all network sizes, both populations, with the range across replicates |
| supp_table_scenarios.csv | The measurement and stationarity scenarios, both populations |
| supp_table_weight_models.csv | The alternative preference models, both populations |
| table2_substitutability.csv | Full model-by-model accuracy grid |

Prepared 2026-07-31.
