## Supplementary material for "Targeting anaemia without measuring it: surrogate prediction, district decision uncertainty and the value of repeat measurement in India": STROBE reporting guidelines

### STROBE reporting checklist

**Study design:** Cross-sectional secondary analysis of published survey estimates and de-identified public-use microdata, with a decision-analytic simulation built on the resulting posterior. The unit of analysis is the district. Items framed around individual participants are mapped to their district-level equivalent, and this is noted where it applies.

Completed against the STROBE statement for cross-sectional studies.

| **Item** | **Recommendation** | **Where addressed** |
| --- | --- | --- |
| **1. Title and abstract** | (a) Indicate the study's design with a commonly used term in the title or the abstract. (b) Provide in the abstract an informative and balanced summary of what was done and what was found. | Title names the analysis rather than a causal claim; the abstract is structured under the journal's four headings and states the design, the data, the analysis, the findings and the conditions under which the decision-analytic figures hold. |
| **2. Background/rationale** | Explain the scientific background and rationale for the investigation being reported. | Introduction, paragraphs 1-4: the change in how India measures anaemia, and why it may or may not matter for decisions. |
| **3. Objectives** | State specific objectives, including any prespecified hypotheses. | Introduction, final paragraph: three explicit questions. Departures from the analysis protocol are declared in the Methods, Step 2, and in the amendment log. |
| **4. Study design** | Present key elements of study design early in the paper. | Methods, opening paragraph and 'Study design, setting and data sources'; the estimand of Step 4 is stated explicitly in that step. |
| **5. Setting** | Describe the setting, locations, and relevant dates, including periods of recruitment, exposure, follow-up, and data collection. | Methods: 707 districts of India, NFHS-5 (2019-21); NFHS-6 fact sheets released 29 May 2026; programme guidelines 29 June 2026. |
| **6. Participants** | Give the eligibility criteria, and the sources and methods of selection of participants. | Methods: the unit of analysis is the district. Two co-primary populations defined by age and haemoglobin threshold; pregnant women excluded with the reason given. |
| **7. Variables** | Clearly define all outcomes, exposures, predictors, potential confounders, and effect modifiers. Give diagnostic criteria, if applicable. | Methods, Steps 2 and 3; Table Ib lists all ten criteria with their availability by survey round. Anaemia thresholds stated explicitly, with an alternative definition tested in Supplement S5. |
| **8. Data sources/measurement** | For each variable of interest, give sources of data and details of methods of assessment. Describe comparability of assessment methods if there is more than one group. | Methods, three numbered sources; agreement between published and recomputed values reported with limits of agreement in Supplement S1.2. The non-comparability of capillary and venous platforms is stated in the Methods and tested in Supplement S4.5. |
| **9. Bias** | Describe any efforts to address potential sources of bias. | No cross-round anaemia comparison; split-sample scoring of selection rules with the ordering statistic computed inside the design half; held-out-State validation; outcome sampling error removed before prediction error is compared with measurement error; matched preference draws across information sets; confounding by indication discussed. |
| **10. Study size** | Explain how the study size was arrived at. | All 707 districts with a published estimate; no sampling of districts. The surrogate analysis is complete case on 693 districts, with the exclusions itemised in the Table I footnote and Supplement S2.2. |
| **11. Quantitative variables** | Explain how quantitative variables were handled in the analyses. If applicable, describe which groupings were chosen and why. | Criteria entered as deficits and modelled on the logit scale; priority reported as three classes with stated thresholds, and the thresholds varied in Supplement S3.4. |
| **12. Statistical methods** | (a) Describe all statistical methods, including those used to control for confounding. (b) Describe any methods used to examine subgroups and interactions. (c) Explain how missing data were addressed. (d) Describe any sensitivity analyses. | Methods, Steps 1-4, with full detail in Supplement S1-S5. Missing criteria drawn from the district's State; missing predictors handled by complete case with the pattern reported. Sensitivity analyses: prior scale, class boundaries, four preference models, decision size 5-20%, platform bias, temporal drift, achieved sample, diagnostic threshold, singleton-stratum variance, and two validation schemes. |
| **13. Participants (flow)** | (a) Report numbers of individuals at each stage of the study. (b) Give reasons for non-participation at each stage. (c) Consider use of a flow diagram. | Not applicable at individual level. District coverage and measured sample sizes reported in the Results and Table Ia; 1 685 994 person-records stated in the Methods. |
| **14. Descriptive data** | (a) Give characteristics of study participants and information on exposures and potential confounders. (b) Indicate the number of participants with missing data for each variable of interest. | Table Ia: prevalence, range, sample size, design effect, the distribution of standard errors and credible interval width by population. Missing values itemised in the Table I footnote and Supplement S2.2. |
| **15. Outcome data** | Report numbers of outcome events or summary measures. | Results, first subsection; Table Ia; per-district values in supp_table_district_results.csv. |
| **16. Main results** | (a) Give unadjusted and confounder-adjusted estimates and their precision. (b) Report category boundaries when continuous variables were categorized. (c) If relevant, consider translating estimates of relative risk into absolute risk. | Results, all subsections; Tables I-II; Figures 1-3. Associations reported with 95% confidence intervals and Holm-adjusted p values; priority class boundaries stated; decision-analytic quantities reported with the range across replicates. |
| **17. Other analyses** | Report other analyses done - eg analyses of subgroups and interactions, and sensitivity analyses. | Abandoned efficiency model recorded as a negative result (Supplement S7); all sensitivity analyses listed under item 12 and reported in Supplement S1.3, S3.4-S3.5, S4.4-S4.5 and S5. |
| **18. Key results** | Summarise key results with reference to study objectives. | Discussion, opening paragraph, mapped to the three questions posed in the Introduction. |
| **19. Limitations** | Discuss limitations of the study, taking into account sources of potential bias or imprecision. Discuss both direction and magnitude of any potential bias. | Discussion, second and third paragraphs: what the decision analysis does and does not estimate, age of the data, ecological design, criterion uncertainty propagated for anaemia only, exploratory status of Step 2, absent population denominators, differing district frames, and the direction each limitation pushes the result. |
| **20. Interpretation** | Give a cautious overall interpretation of results considering objectives, limitations, multiplicity of analyses, results from similar studies, and other relevant evidence. | Discussion: an explicit statement that the findings are not evidence that supplementation is ineffective, that no figure here is a realised cost, and that the multicriteria and burden-only analyses answer different questions. Multiplicity handled by Holm adjustment in Supplement S2.5. |
| **21. Generalisability** | Discuss the generalisability (external validity) of the study results. | Discussion, final paragraph: findings apply to district-level targeting in India under the stated measurement model; the held-out-State analysis addresses transfer to unobserved regions and the scenario analysis addresses transfer to a different platform and a later period. |
| **22. Funding** | Give the source of funding and the role of the funders for the present study and, if applicable, for the original study on which the present article is based. | Declarations: none. The underlying surveys are funded by the Government of India and its partners, and had no role in this analysis. |

An explanation and elaboration article discusses each checklist item and gives methodological background and published examples of transparent reporting. The STROBE checklist is distributed under a Creative Commons Attribution licence.

Prepared 2026-07-31.
